# Breakthrough infections with the omicron and delta variants of SARS-CoV-2 result in similar re-activation of vaccine-induced immunity

**DOI:** 10.1101/2022.01.27.22269895

**Authors:** Arne Søraas, Gunnveig Grødeland, Beathe Kiland Granerud, Thor Ueland, Andreas Lind, Børre Fevang, Sarah Murphy, Camilla Huse, Anders Benteson Nygaard, Anne Katrine Steffensen, Huda al-Baldawi, Mona Holberg-Petersen, Lise Lima Andresen, Camilla Ågnes, Trine Ranheim, Ylva Schanke, Mette Istre, John Arne Dahl, Adity Chopra, Susanne Dudman, Mari Kaarbø, Jan Terje Andersen, Eline Benno Vaage, Trung The Tran, John Torgils Vaage, Annika E. Michelsen, Fredrik Müller, Pål Aukrust, Bente Halvorsen, Tuva Børresdatter Dahl, Jan Cato Holter, Fridtjof Lund-Johansen

## Abstract

**Background:** The Omicron variant of SARS-CoV-2 is now overtaking the Delta variant in many countries. Results showing that sera from double vaccinated individuals have minimal neutralizing activity against Omicron may indicate that the higher rate of transmission is due to evasion from vaccine-induced immunity. However, there is little information about activation of recall responses to Omicron in vaccinated individuals.

**Methods:** We measured inflammatory mediators, antibodies to the SARS-CoV-2 spike and nucleocapsid proteins, and spike peptide-induced release of interferon gamma in whole blood in 51 vaccinated individuals infected with Omicron, in 14 infected with Delta, and in 18 healthy controls. The median time points for the first and second samples were 7 and 14 days after symptom onset, respectively.

**Findings:** Infection with Omicron or Delta led to a rapid and similar increase in antibodies to the SARS-CoV-2 spike and nucleocapsid proteins and spike peptide-induced interferon gamma in whole blood. Both the Omicron and the Delta infected patients had a mild and transient increase in inflammatory parameters.

**Interpretation:** The results suggest that vaccine-induced immunological memory yields similar coverage for the Omicron and Delta variants.

## INTRODUCTION

The SARS-CoV-2 Omicron (B.1.1.529) variant is now dominating the pandemic in many countries. The virus harbors up to 36 mutations in the spike protein, which is the target of vaccine-induced neutralizing antibodies (https://covdb.stanford.edu/page/mutation-viewer/#Omicron). Sera from individuals who have received two vaccine doses appear to have little or no neutralizing activity to Omicron, and early results from South Africa and the UK suggest that vaccine effectiveness against symptomatic disease with Omicron is significantly lower than with the Delta (B.1.617.2) variant.^1-7^ There are now concerns that standard vaccine dose regimens may not provide sufficient protection, and that booster doses are necessary. While it seems possible that Omicron evades immunological memory established after vaccination, there is currently little knowledge about immune responses to Omicron infection. To assess re-activation of humoral immunological memory, we measured the early SARS-CoV-2 induced immune responses in twice vaccinated individuals infected with Omicron during an outbreak in Oslo in November 2021.^8^ Double-vaccinated individuals infected with the Delta variant served as controls. We present data on viral load in nasopharynx, the initial general inflammatory response, antibodies to protein- and peptide antigens derived from SARS-CoV-2 and seasonal coronaviruses, and activation of spike-specific T cells as assessed by interferon (IFN)-γ in blood upon exposure to spike peptides.

## METHODS

Between November 30 and December 11, 2021, adults (≥18 years old) in Oslo and the surrounding county Viken with positive SARS-CoV-2 RT-PCR test, with or without positive Omicron or Delta variant PCR on oro-nasopharyngeal specimens (i.e., suspected Omicron and verified Delta variant cases, respectively), and symptomatic household members of suspected Omicron cases were consecutively recruited to a prospective cohort study [a joint venture between The Norwegian Corona Cohort (NCT04320732) and the Norwegian SARS-CoV-2 study (NCT04381819)].

Clinical data and samples (nasopharyngeal swabs and blood samples) were collected at the earliest time point after diagnosis (i.e., inclusion) and at 1 week of follow-up. Data were collected using electronic questionnaires based on an adapted version of The International Severe Acute Respiratory and Emerging Infection Consortium (ISARIC) Tier 1 Initial Freestanding follow up survey.^9^ The samples were taken at the patients` home by an ambulant team, or at an outpatient clinic at Oslo University Hospital (OUH). Data on SARS-CoV-2 vaccination status were obtained from the Norwegian national mandatory registry on vaccination (SYSVAK).

Initially, a total of 41 Omicron suspected and 18 verified Delta variant cases were included in the study. Among 16 household members of Omicron suspected cases, 11 were considered Omicron suspected cases and included in the study according to the test criteria, thus ending with 52 cases in the Omicron group and 18 in the Delta group (**Table 1**). All suspected Omicron cases were verified by whole genome sequencing.^10^ Household members who tested negative were included in the analysis of healthy controls (**Supplemental Table 1**).

**Table 1.** Demographics, clinical characteristics, and laboratory findings at inclusion

|  | Omicron, n=52 | Delta, n=18 | p-value |
| --- | --- | --- | --- |
| Age, years | 38.9±12.3 | 39.9±8 | 0.73 |
| Male gender | 25 (48.1%) | 7 (38.9%) | 0.50 |
| BMI | 23.6±4 | 24.7±3.8 | 0.33 |
| Previous COVID-19 infection | 5 (10%) | 0 (0%) | 0.20 |
| Bedridden, (n=no/1-6d/7-13d) | (34/16/2) | (9/8/1) | 0.51 |
| Comorbidity |  |  |  |
| Cardiac | 0 (0%) | 0 (0%) | 1.00 |
| Hypertension | 0 (0%) | 1 (5.9%) | 0.25 |
| COPD | 1 (2%) | 0 (0%) | 0.74 |
| Asthma | 3 (5.8%) | 1 (5.9%) | 0.69 |
| Diabetic | 0 (0%) | 1 (5.6%) | 0.27 |
| Cancer | 0 (0%) | 0 (0%) | 1.00 |
| Other disease | 5 (10.2%) | 3 (16.7%) | 0.37 |
| Medication count (n=0/1/≥2) | (35/14/3) | (12/3/3) | 0.10 |
| Immuno-suppression | 0 (0%) | 0 (0%) |  |
| Smoke (n=no/yes/previuos) | (41/7/2) | (14 /3/1) | 0.92 |
| Poor fitness | 16 (30.8%) | 8 (44.4%) | 0.22 |
| Days from symptom onset to Inclusion | 6±3 | 7±3 | 0.60 |
| Days from symptom onset to follow up sample | 15±3 | 14±4 | 0.23 |
| Vaccinations, (n=0/1/2/3) | (1/3/47/1) | (0/0/16/2) | 0.53 |
| Biochemistry |  |  |  |
| Hemoglobin, g/dL | 14.5 (13.5, 15.1) | 14.0 (13.1, 14.5) | 0.073 |
| WBC, x10<sup>9</sup>/L | 5.2 (4.3, 5.9) | 5.3 (4.5, 7.0) | 0.63 |
| eGFR, mL/min/1.73m<sup>2</sup> | 113 (103, 119) | 108 (100, 114) | 0.10 |
| Ferritin, µg/L | 119 (66, 221) | 159 (72, 201) | 0.80 |
| CRP, mg/L | 1.8 (0.7, 4.9) | 1.9 (1.0, 4.4) | 0.93 |
| D-dimer, mg/L FEU | 0.23 (0.18, 0.32) | 0.23 (0.19, 0.31) | 0.61 |

Data are n (%), mean ± SD or median (IQR). Abbreviations: BMI, Body mass index; COPD, chronic obstructive pulmonary disease; WBC, white blood cells; eGFR, estimated glomeruli filtration rate; CRP, C-reactive Protein; D-dimer; FEU, fibrinogen equivalent units.

Reference samples were also collected in the same period of time from 14 age and sex-matched vaccinated healthy controls who were recruited at the Research Institute of Internal - Medicine, OUH (**supplemental Table 1**).

Informed consent was obtained from all individuals. The study was approved by the Regional Committee for Medical and Health Research Ethics in South-Eastern Norway (reference numbers 124170 and 106624).

### Blood sampling protocol

Blood for routine biochemistry, SARS-CoV-2 specific antibodies, interferon gamma (IFN-γ) release assay (IGRA), and markers of immune activation and inflammation was taken at baseline within a median of 6 days after symptom onset (Table 1), and at 1 week follow-up (4-6 days from inclusion). Samples were processed within 1.5 hour, or centrifuged at 2000*g* for 20 minutes at 4ºC (EDTA plasma; serum: 15 minutes at room temperature) and stored at - 80ºC until further analysis.

### Biochemical analyses

Routine blood biochemistry including C-reactive protein (CRP), ferritin, lactate dehydrogenase (LDH), haemoglobin, creatinine, alanine aminotransferase (ALT), aspartate aminotransferase (AST), fibrinogen, procalcitonin (PCT), D-dimer, platelet count, total white blood cell (WBC) count, monocyte, neutrophil and lymphocyte count, immunoglobulin (Ig), troponin T (TnT), and N terminal pro-brain-natriuretic peptide (NT-proBNP) were analysed at Laboratory for Medical Biochemistry at OUH Rikshospitalet, Oslo, Norway. Plasma levels of soluble (s) CD14, sCD163, sCD25, soluble T cell immunoglobulin mucin domain-3 (sTIM-3), myeoloperoxidase (MPO), lipopolysaccharide (LPS) binding protein (LBP), IFNγ-induced protein (IP-10), pentraxin-3 (PTX-3), growth differentiation factor 15 (GDF-15), matrix metallopeptidase 9 (MMP-9), and S100 calcium-binding protein A12 (S100A12) were measured in duplicate by enzyme immunoassays (EIA) using commercially available antibodies (R&D Systems, Minneapolis, MN) in a 384 format using a combination of a SELMA (Jena, Germany) pipetting robot and a BioTek (Winooski, VT) dispenser/washer. The terminal complement complex TCC was measured with the same setup using a monoclonal antibody aE11 reacting with a neoepitope exposed in C9 when incorporated in TCC, in a modified version of the method described by Mollnes et al.^11,12^ A similar setup was used for von Willebrand factor (vWF) with antibodies from Dako Cytomation (Glostrup, Denmark) using parallel diluted human plasma as standard curve. Absorption was read at 450 nm with wavelength correction set to 540 nm using an EIA plate reader (BioTek). Samples from all patients and controls were run on the same 384-well plate and intra-assay coefficient of variation was <10%.

### Antibody measurement

Antibodies to the nucleocapsid and RBD proteins derived from wild-type SARS-CoV-2 were measured with using a slightly modified version of a bead-based assay described previously.^13^ Briefly, neutravidin-coupled polymer beads with fluorescent barcodes and biotinylated virus proteins were incubated for 1h with serum diluted 1:1000 or 1:10.000 at 22ºC. SARS-CoV-2 RBD ad full-length spike were obtained from Florian Krammer and Ian McLellan, respectively.^14,15^ Spike proteins from seasonal coronaviruses were obtained from Ulrich Rothbauer.^16^ Peptide antigens from SARS-CoV-2 were described previously.^17^ The beads were washed twice with PBS containing 1% Tween 20 (PBT), and labelled with R-Phycoerythrin-conjugated anti-human IgG Fc or anti-human IgA Fc (Jackson Immunoresearch). Next, the beads were analyzed by flow cytometry. Antibody levels were measured as the R-Phycoerythrin median fluorescence intensity (MFI) of beads coupled with virus proteins divided by the signal measured for beads coupled with neutravidin only. A serum sample measured to contain 53000 Binding Antibody Units (BAU) with the Roche Elecsys assay was used to generate a standard curve to convert relative MFI values for IgG to BAU.

### Measurement of neutralizing antibodies

Vero E6 cells were added into 96-well plates (Costar 3595, Corning Incorporated) in 1×10^4^ cells/well. The following day, sera were inactivated at 56°C for 30 min, and titrated amounts mixed with TCID100 of SARS-CoV-2 virus (Human 2019-nCOV strain 2019-mCoV/Italy-INM1) in quadruples. Following a one hour incubation, the mixtures were added to the cells and incubated for 4 days at 37ºC in a 5% CO_2_ humidified atmosphere. Next, the plates were washed with PBS and fixed with acetone/PBS for 30 min. The plates were air dried and incubated with rabbit anti-SARS-CoV-2 nucleocapsid antibody (cat. 40143-R004, Sino Biological) over night at 4°C. Plates were now incubated with horseradish peroxidase (HRP)-conjugated anti-rabbit IgG Fc antibody (cat. SSA003, Sino Biological) for 1 hour at room temperature, developed with TMB Substrate Solution (cat. N301, ThermoFisher), and read with an EnVision 2104 Multilabel Reader (Perkin Elmer). The assay has been validated by comparison with other laboratories.^18^

### Detection of SARS-CoV-2 T cell response by IFNγ-release assay (IGRA)

The detection of SARS-CoV-2 CD4 and CD4/CD8 spike T cell responses was done using QuantiFERON SARS-CoV-2 IGRA (Qiagen, Germany) according to the manufacturer’s instructions.^19^ After tube processing and incubation, analysis was done on a LIAISON XL fully automated chemiluminescence analyzer (Diasorin, Italy). Pre-validated cut-offs for antigen specific responses were defined to be ≥0.101 IU/mL and ≥0.145 IU/mL for CD4 (Ag1) and CD4/CD8 (Ag2) T cells, respectively, after subtraction of background (Nil). Results < 0.03 IU/mL was considered a non-response.

### Viral load quantification in nasopharyngeal swabs

Nasopharyngeal swabs were frozen at -20ºC upon arrival, thawed, and inactivated at 56ºC for 30 min. Bacteriophage MS2 (Roche, Switzerland) was added to samples as extraction/inhibition control. Nasopharynx samples (200 uL) were extracted on the MagNaPure 96 system, using MagNaPure 96 DNA and Viral NA Small Volume kit (Roche, Switzerland) in combination with the universal pathogen protocol and elution in 50 uL. SARS-CoV-2 RNA real-time reverse transcriptase PCR targeting the viral envelope gene was done as previously described, on the LightCycler 480 instrument (Roche, Switzerland).^20^ Samples with Cp-values >35 were considered negative. Cellular quantification in nasopharyngeal samples was assessed using the CELL Control r-gene kit targeting the HPRT1 gene (bioMérieux, France) according to the manufacturer’s instructions. Viral quantification was performed using dilution series of standards calibrated against the First WHO International Standard for SARS-CoV-2 (reference standard 20/146, NIBSC, England). Viral load in nasopharynx samples was determined as virus RNA copies per 1000 human cells.

### Statistical analysis

Differences in categorical demographics, routine biochemistry according to reference limits and symptom scores were analyzed with chi-square or Fisher’s exact test. Differences in age and BMI were compared using students T-test. Biochemical data and markers of immune activation and inflammation were compared with non-parametric tests; Kruskal-Wallis a priori if >2 groups (i.e. when the control group was included) and Mann-Whitney when two groups. When adjustment for covariates was necessary, ANCOVA was used (e.g., when assessing virus load between Omicron and Delta using symptom days as covariate). We used a linear mixed model to assess the association between virus load (log10 transformed) and symptom days with a random intercept by subject to control for repeated measures. Significance of difference in antibody levels and interferon gamma release was tested using Wilcoxon Signed-Rank Test for paired samples or Mann-Whitney with help of the Excel plugin Realstats. P-values are two-sided and considered significant when <0.05 (**Supplemental Table 1**).

## RESULTS

### Cohort characteristics

Demographics and clinical characteristics at inclusion of the COVID-19 patients infected with Omicron (n=52) or Delta (n=18) variants is shown in **Table 1**. Overall, the characteristics, including symptom duration before diagnosis, were similar between the two groups. Median age was approximately 40 years, and 80% were <50 years of age. There were few co-morbidities and no patients reported the use of immuno-suppressive drugs. All but one had received at least one dose of an mRNA COVID-19 vaccine, and 65 had received ≥2 doses (**Table 1**). One individual with Omicron and two with Delta had received a third dose. Five patients with Omicron, but none with Delta, reported previous COVID-19. Information about vaccine status was obtained for 13 out of 14 healthy controls. All had received two doses, one had received a booster dose, and one reported previous COVID-19 (**Supplemental Table 1**).

### Symptomatology

Most patients experienced mild symptoms with nasal symptoms, cough being the most frequent (67-78%) (**Table 2**). Around 50% experienced fever and around 5% high fever (>39ºC). None of the patients needed hospitalization. There were few differences in symptoms between Omicron and Delta infected patients. However, while smell/taste symptoms were frequent in Delta patients (72%) they were infrequent in Omicron patients (15%, p<0.001). A larger proportion of Delta patients experienced concentration difficulties (“brain fog”, 33%) compared to Omicron patients (10%, p=0.017).

**Table 2.** Symptoms

|  |  | Omicron,<br>n=52 | Delta, n=18 | p-value |
| --- | --- | --- | --- | --- |
| Total symptoms reported |  | 50 (98%) | 17 (94.4%) | 0.46 |
| Fever |  | 26 (50%) | 8 (44.4%) | 0.68 |
| Fever >39 degrees |  | 2 (3.8%) | 1 (5.6%) | 0.60 |
| Dyspnea |  | 13 (25%) | 6 (33.3%) | 0.49 |
| Cough |  | 39 (75%) | 12 (66.7%) | 0.49 |
| Fatigue |  | 29 (55.8%) | 12 (66.7%) | 0.42 |
| Muscular pain |  | 27 (51.9%) | 9 (50%) | 0.88 |
| Sore throat |  | 33 (63.5%) | 7 (38.9%) | 0.069 |
| Changed sense of smell and taste |  | 8 (15.4%) | 13 (72.2%) | <0.001 |
| Nasal symptoms |  | 38 (73.1%) | 14 (77.8%) | 0.69 |
| Headache |  | 37 (71.2%) | 10 (55.6%) | 0.23 |
| Abdominal symptoms |  | 7 (13.5%) | 4 (22.2%) | 0.38 |
| Memory problems |  | 0 (0%) | 0 (0%) | 1.00 |
| Problems concentrating |  | 5 (9.6%) | 6 (33.3%) | 0.017 |
| Disorientated |  | 0 (0%) | 0 (0%) | 1.00 |
| Feeling dizzy |  | 15 (28.8%) | 5 (27.8%) | 0.93 |
| Other symptoms |  | 4 (7.7%) | 1 (5.6%) | 0.62 |
| Health assessment | good | 18 (34.6%) | 6 (33.3%) | 0.62 |
|  | fair | 27 (51.9%) | 11 (61.1%) |  |
|  | poor | 7 (13.5%) | 1 (5.6%) |  |
| Symptom duration, days |  | 5±2 | 5±2 | 0.87 |
| Average # symptoms |  | 5±3 | 6±3 | 0.82 |

Data are shown as n (%) or mean ± SD.
The table is based on self-report of symptoms through a questionnaire and lists all symptoms reported from disease onset to inclusion in the study.
SD=Standard deviation

### Viral load in nasopharynx

Using a mixed model adjusting for repeated measures, viral load in nasopharynx was inversely correlated with time since symptom debut (t=-3.9, p<0.001) (**Figure 1A**), showing no differences in the slopes between Omicron and Delta variants. However, after adjusting for time since symptom onset, viral load was significantly higher in patients infected with Omicron at 1 week follow-up, compared with those infected with the Delta variant, reflecting that all Delta infected patients had undetectable virus (**Figure 1B**). Importantly, however, as many as 80% of Omicron patients had undetectable virus levels at this time point.

**Figure 1:**
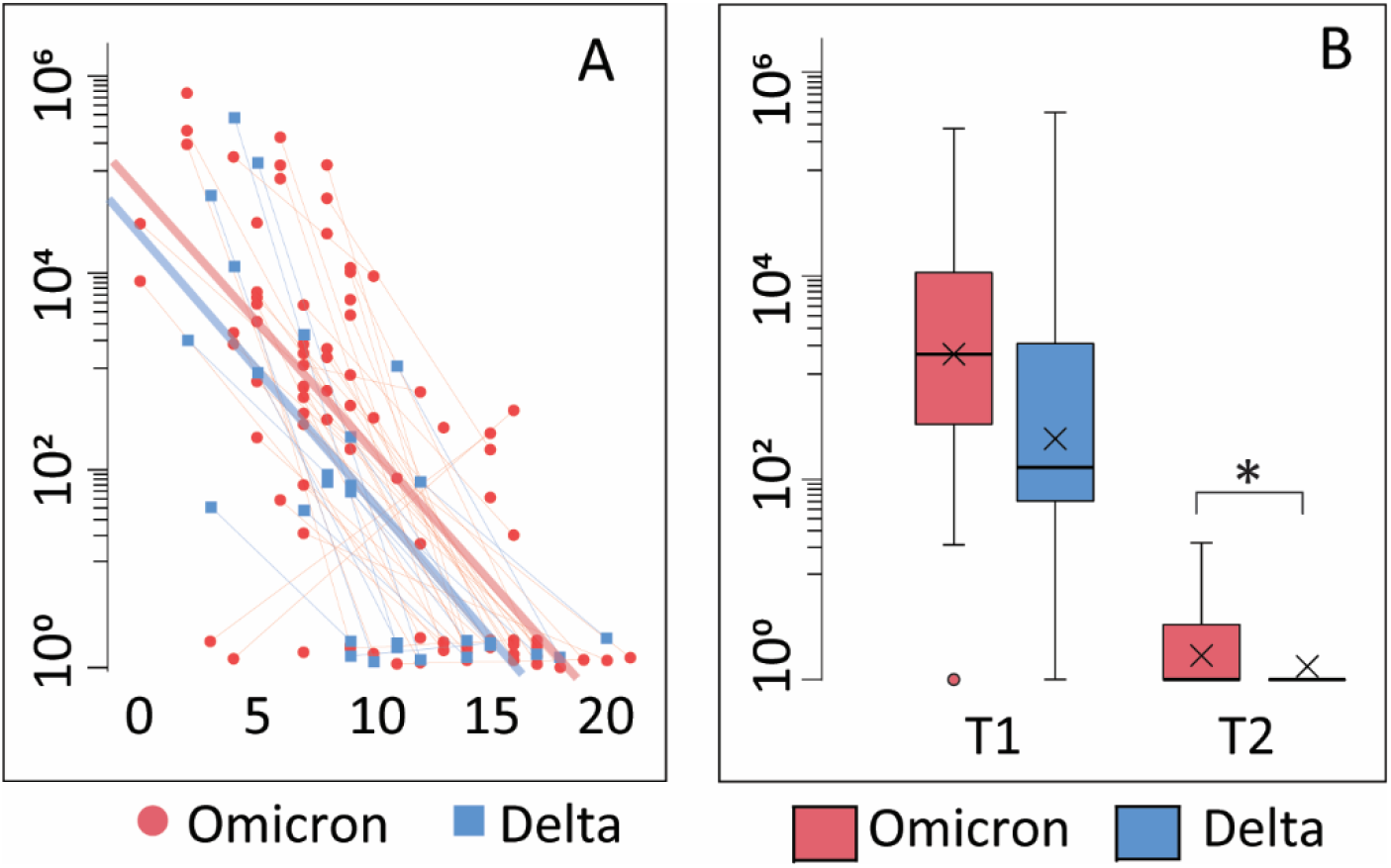
Viral load. A: Correlation between viral load and days after symptom onset during the observation period. Thin blue/red lines represent paired samples while thick blue/red lines represent regression curves for the whole group (Omicron or Delta). B: Viral load shown as Tukey-plots at inclusion (T1) and one week follow-up (T2) according to infection with Omicron or Delta. *p<0·05 adjusting for symptom days.

### Routine biochemistry

Most measures, including hematology, liver enzymes, kidney function, and cardiac markers, were normal, and the 0-15% outside reference limits were only marginally elevated (**Supplemental Table 2**). Between 25-30% had elevated CRP and fibrinogen at inclusion, which normalized at 1 week. There were no differences in routine biochemistry between Omicron and Delta patients, except higher IgA levels in the Delta group at inclusion and one week, but with no difference in the proportion of patients with IgA outside reference levels between the groups. In the patient group as a whole, those with smell/taste symptoms had higher IgA levels than those without these symptoms at inclusion (median 2.4 g/L vs. 1.9 g/L, p=0.021) with a similar trend at 1-week (2.2 g/L vs. 1.8 g/L, p=0.054).

### Immune activation and inflammation

We measured plasma levels for a wide range of markers of inflammation and immune activation at inclusion and one-week follow up, according to Omicron- or Delta infection and in healthy controls (**Supplemental Table 3**). A few markers were increased at inclusion as compared to healthy controls. Thus, the monocyte/macrophage activation markers sCD14 and sCD163, the acute phase markers LBP and PTX-3, IP-10 with activating effects on T cells, and the fibrosis marker GDF-15 were all elevated compared with controls, but with no differences between Omicron and Delta (**Supplemental Table 3**). These markers returned to levels comparable to controls after one week, except PTX-3 that remained elevated in patients with Delta infection after one week, and GDF-15 that remained elevated in both groups (**Supplemental Table 3**).

### Omicron and Delta lead to a similar increase in T-cell release of IFN-γ

Whole blood IFN-γ release assays were used for indirect detection of spike-reactive T-cells. Levels of secreted IFN-γ was similar in individuals infected with Omicron and Delta, and higher than observed in healthy vaccinated controls (**Fig. 2A**). There were no time-dependent changes in INF-γ release between inclusion and at one week follow-up. Collectively, these results suggest that memory T-cells were expanded at a very early stage during infection.

**Figure 2:**
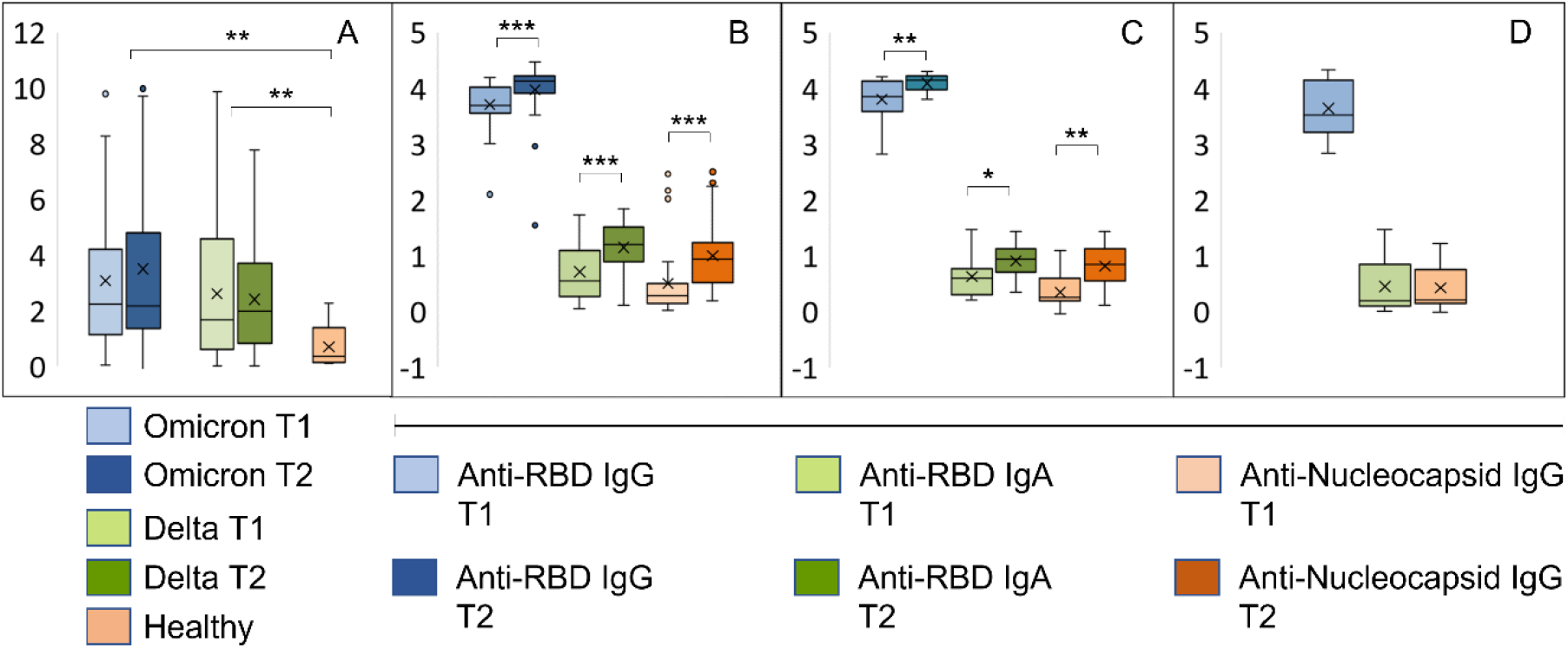
Early immune responses in individuals infected with Omicron or Delta. A: Results from whole blood IFNγ release assay (IGRA, IU/ml) in samples harvested from the same individuals with an interval of 8-10 days. T1: inclusion, T2: one week follow up. B-D: The bar graphs show relative levels (log 10) of antibodies to RBD and the nucleocapsid protein in samples described under A. B: Individuals with confirmed Omicron infection, C: Delta infection, D: vaccinated individuals with no history of SARS-CoV-2 infection. ***p<10^−5^, **p<10^−3^, *p<10^−2^. Source data are found in Supplemental Table 1.

### Omicron and Delta lead to a similar increase in the levels of antibodies to RBD in vaccinated individuals

All but one study participant was positive for anti-RBD antibodies, and the majority had titers higher than 4000 BAU (**Figure 2B-C, Supplemental Table 1**). A total of 69 sera were tested for neutralizing activity against vaccine ancestral SARS-CoV-2. The median titer was 450 (range 28-640), and the correlation with BAU was 0·77 (**Supplemental Table 1**). Infection with Omicron or Delta led to a modest and similar time-dependent increase in anti-RBD titers (**Figure 2B-C**). The two groups also had a similar increase in IgA antibodies to RBD and IgG antibodies to nucleocapsid (**Figure 2B-C**). High levels of IgG antibodies to RBD were also found in samples from healthy vaccinated controls, whereas anti-RBD IgA and anti-nucleocapsid IgG were nearly undetectable (**Figure 2D**).

The dot plots in **Figure 3 A-L** show time-resolved data at the individual level. While most infected individuals had cleared the viral infection by day 12, virus could be detected also longer in a higher number of individuals infected with Omicron as compared to Delta (**Figure 3A and G**). Antibody responses against RBD (Wuhan and Omicron) and nucleocapsid, as well as cellular immune responses were, however, similar between the groups (**Figure 3 B-L**). Five study participants had high levels of antibodies to the nucleocapsid protein in both samples (**Figure 3C**). These all had self-reported previous COVID-19 (**Supplemental Table 1**).

**Figure 3:**
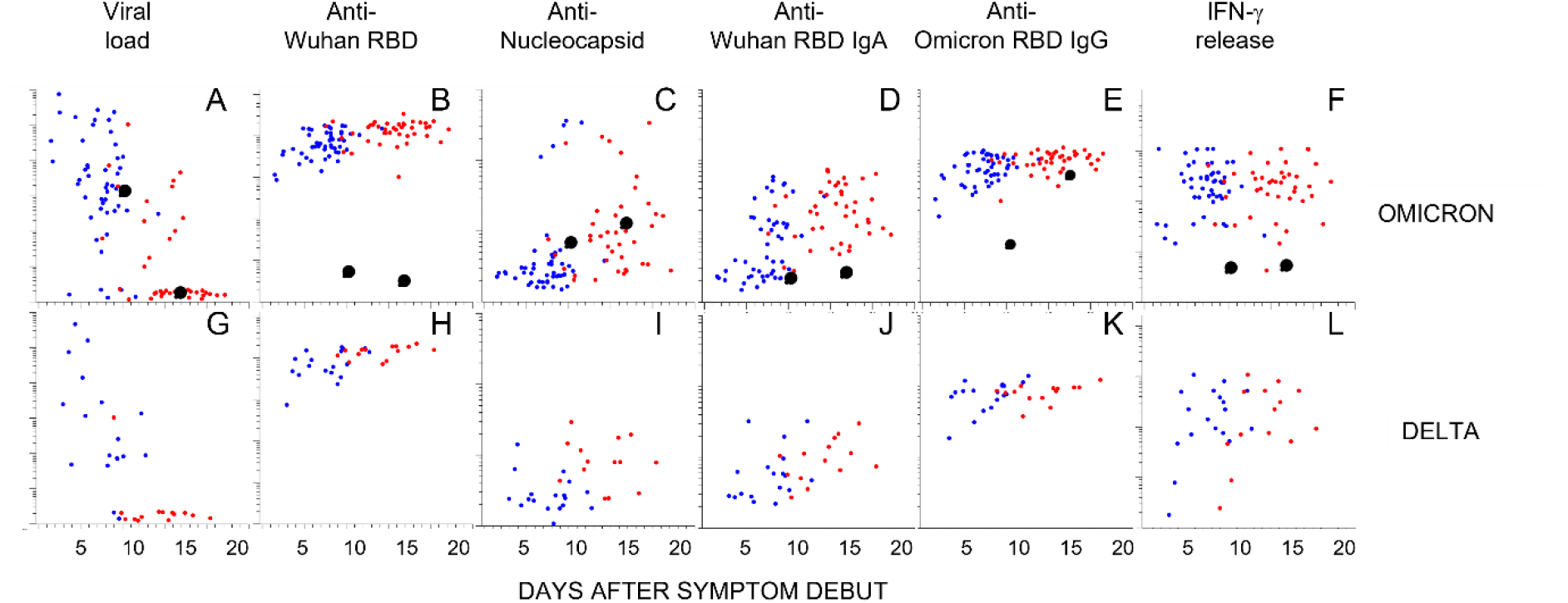
Kinetics of antibody responses in individuals infected with Omicron or Delta. The scatter plots show individual data after infection with Omicron (A-F) or Delta (G-L) plotted at days after symptoms onset. A and G: Viral load in nasopharyngeal swabs. B-E and H-K: Relative levels of antibodies to indicated antigen (y-axis, log10). F and L: T-cell responses by IFNγ release assay. Blue and red dots indicate the first and second sample obtained at 4-14 days intervals, respectively. Enlarged black dots indicate samples from an unvaccinated individual obtained at 9 or 16 days after symptom onset. Source data are found in Supplemental Table 1.

One participant infected with Omicron was unvaccinated and had no previous history of COVID-19 (**Figure 3, black enlarged dots**). This individual cleared the virus at day 16 (**Figure 3A**). Antibodies to Wuhan RBD, nucleocapsid, or IFN-γ release, were not detected, but reactivity with Omicron RBD was detected at day 16 (**Figure 3B-F**). Interestingly, there were high levels of antibodies to full length Spike protein (Wuhan) in the first sample (**Figure 4A**). There was also a high level of antibodies to the S2 domain of the seasonal coronavirus OC43, and to two peptides that are conserved in the S2 domains of SARS-CoV-2 and OC43 (**Figure 4A-B**). However, there were no elevation of antibodies to the S1 domains of spike from OC43 or HKU1 or to influenza or rhinovirus (**Figure 4D-E**).

**Figure 4:**
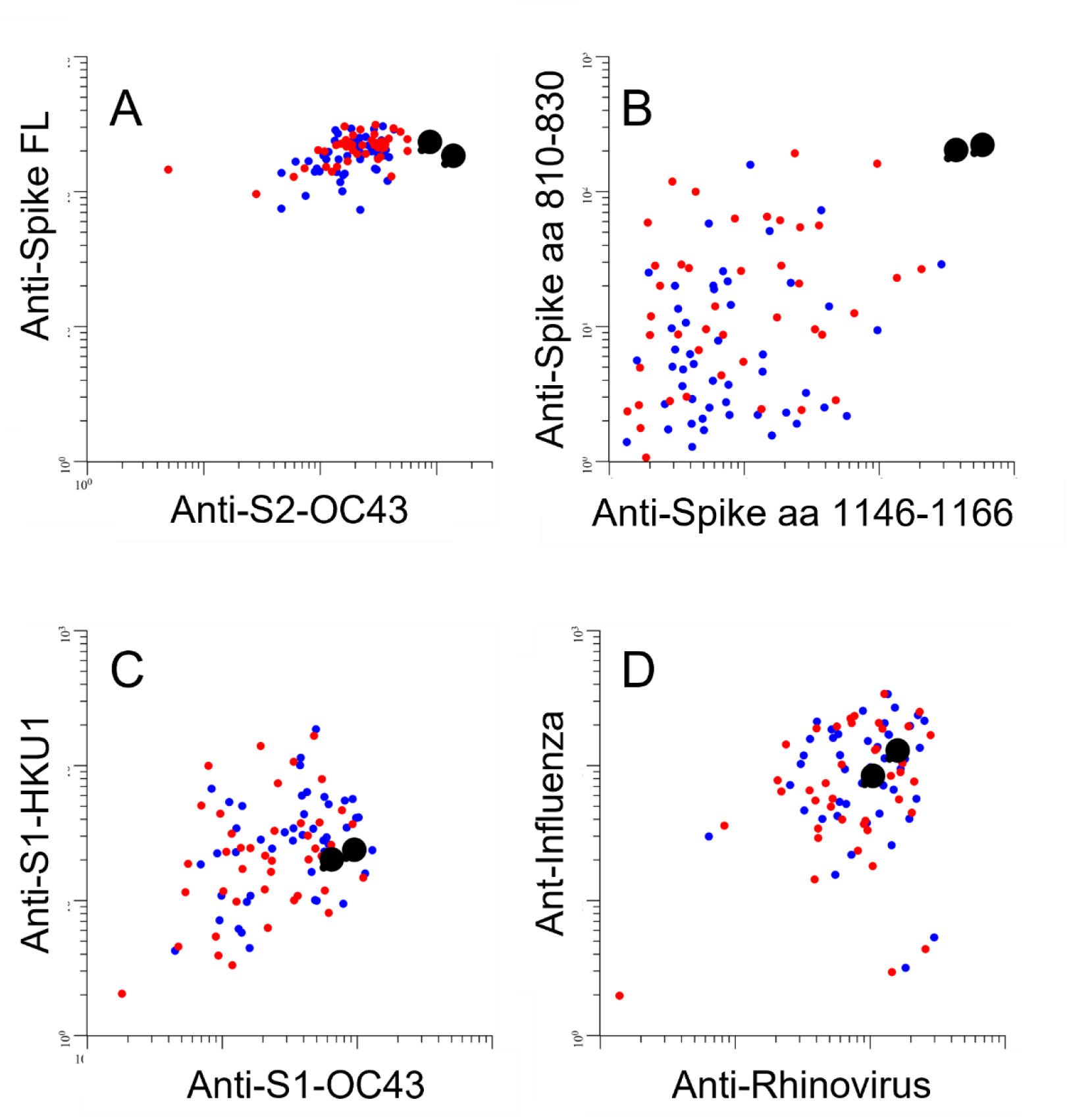
Levels of antibodies to spike proteins from SARS-CoV-2 and seasonal coronaviruses in individuals infected with Omicron. The scatter plots show relative levels of IgG antibodies to indicated antigen in individuals infected with Omicron [spike-full length protein (Spike FL) from the SARS-CoV-2 Wuhan strain. S2-OC43: S2 domain of seasonal coronavirus OC43, S1 HKU1/OC43: S1 domains from seasonal coronaviruses HKU1 and OC43, respectively. Spike aa 810-830 and aa 1146-1166: linear peptide antigens that are conserved in the S2 domains of spike proteins in SARS-CoV-2 and seasonal coronaviruses OC43 and HKU1.^17^] Enlarged black dots correspond to unvaccinated individual (see Fig. 3).

## DISCUSSION

We have measured immune responses in vaccinated individuals at two time points during the first two weeks after infection with the Omicron or Delta variants of SARS-CoV-2. Since the majority had received two doses of mRNA vaccines more than three months prior to infection, we expected that infection with Delta would mimic a booster dose and lead to a large increase in spike-reactive antibodies. Serum from individuals who had received two vaccine doses were recently reported to have greatly reduced neutralizing activity against Omicron.^1-4,7^ We therefore also expected that infection with this variant would lead to weak humoral recall responses as compared to Delta. The results obtained here show that infection with both variants were associated with mild symptoms, low levels of inflammatory mediators, and a modest increase in levels of antibodies to RBD. Results obtained by measuring IFN-γ release in whole blood after stimulation with peptides from the vaccine ancestral SARS-CoV-2 spike protein suggested that there was a rapid and similar activation of T-cell mediated immunity after infection with both variants. This was expected from previous results showing conservation of T-cell-mediated immunity.^3,7,21,22^ Collectively, the results suggest that infection with Omicron and Delta variants lead to similar activation of humoral and cellular recall responses in individuals who have received two doses of mRNA COVID-19 vaccines.

At one week after inclusion (i.e., 13 days after symptom debut) all nasopharynx swabs obtained from individuals infected with Delta were negative. However, as many as 80% of the Omicron patients also had undetectable virus levels at this time point. Due to limitations in the cohort size and observation period, one cannot draw firm conclusions about the exact kinetics of virus clearance. Yet, in view of the mild symptoms and rapid clinical recovery it seems likely that the combination of circulating antibodies and recall responses in this cohort of double-vaccinated individuals was sufficient for full recovery. This interpretation is in agreement with results from a study showing that mRNA vaccines elicit highly cross-reactive antibodies to SARS-CoV-2 variants.^22^

The methods used here do not discriminate between primary and recall responses to the SARS-CoV-2 spike protein. Yet, results obtained for antibodies specific for the nucleocapsid protein suggest that the contribution of primary responses was modest. There was a statistically significant increase in anti-nucleocapsid antibodies over time, but except for those with a prior history of COVID-19, the levels remained near the detection limit in most individuals throughout the observation period. The levels of spike peptide-induced IFN-γ release from T-cells were higher than those observed in samples from healthy controls already at 2-10 days after symptom onset. This is well below the time range expected for a primary T-cell response. It therefore is reasonable to conclude that virus clearance was mainly mediated through recall responses.

Results obtained from the single unvaccinated individual infected with Omicron should be interpreted with great caution, but they are nevertheless interesting. The selective IgG reactivity to Omicron RBD at day 16 suggests that infection with this variant does not necessarily lead generation of to antibodies that neutralize other variants. The high levels of antibodies to full-length spike protein from the Wuhan variant at day 9 is likely to be directed to the S2 domain. While we did not have access to pre-infection samples, we have previously shown that antibodies to the full-length spike protein are rarely found in pre-pandemic samples.^13^ It is also worth noting that the unvaccinated study participant with Omicron infection had very high levels of antibodies to the S2 domain of seasonal coronavirus OC43, and to peptide epitopes in S2 that are conserved between SARS-CoV-2 and OC43. The most likely explanation for reactivity to the full-length spike protein at day 9 is therefore a cross-reactive recall response. While this was only observed in a single individual, the rapid clearance of virus may indicate that cross-reactive S2-reactive antibodies developed during infection with seasonal coronaviruses in some cases can be broadly protective.

It is important to note that the cohort consisted of otherwise healthy individuals, the majority younger than 50 years of age, and that the median time since the second vaccine dose was only 3.8 months (**Supplemental Table 1**). While we did not have a true baseline, the median titer of anti-RBD antibodies in the first set of samples was higher than 4000 BAU, which was found to correspond to a neutralization titer of 450 against ancestral vaccine SARS-CoV-2 (**Supplemental Table 1**). Thus, the immune status in this cohort is probably well above the average for people who have received two doses of mRNA vaccines during 2021. Yet, the levels are probably not higher than what is obtained in older individuals after a booster dose. The four participants who had received a booster dose had anti-RBD antibody titers in the range of 8000-16000 BAU (**Supplemental Table 1**).

It is well established that vaccination of COVID-19 convalescents results in strong and lasting immunity. Less is known about the immunity obtained when vaccinated individuals are infected. Here, we show that the levels of antibodies to the nucleocapsid protein remain low during the first two weeks after vaccinated individuals were infected with Delta or Omicron. Longer observation periods are necessary to firmly establish the level of immunity that is obtained. Yet, it seems possible that vaccinated individuals may clear the virus before they mount a strong primary immune response. This possibility should be addressed in studies with longer follow up times.

In conclusion, we show that infection with Omicron and Delta results in similar activation of immune recall responses in individuals who have received two doses of mRNA COVID-19 vaccines. Omicron evasion from humoral immunity may therefore be less severe than indicated from results of *in vitro* virus neutralization assays.

## Data Availability

All data produced in the present study are available upon reasonable request to the authors

## ACKNOWLEDGEMENTS

The authors would like to thank Cathrine Fladeby for providing professional support with virus quantification. We thank Roger Eugen Poliakov Egseth for technical support.

## AUTHOR CONTRIBUTIONS

AS – study design, conceptualization, sample collection, data collection, data curation, interpretation, project administration, funding acquisition, writing original draft, writing review and editing; GG – study design, sample analysis, interpretation, writing review and editing; BKG – sample analysis, validation, interpretation, writing original draft, writing review and editing; TU – sample analysis, data curation, data analysis, interpretation, visualisation, writing original draft, writing review and editing; AL – sample collection, data collection, sample analysis, interpretation, writing original draft, writing review and editing; BF – sample collection, data collection, writing review and editing; SLM – sample collection, writing review and editing; CH – sample collection, writing review and editing; ABN – data collection, data curation, writing review and editing; AKS – sample analysis, validation, writing review and editing; Ha-B – sample analysis, validation, writing review and editing; MH-P – sample analysis, validation, data curation, writing review and editing; LLA – data collection, sample analysis, writing review and editing; CÅ – data collection, writing review and editing; TR – sample analysis, writing review and editing; YS – sample analysis, writing review and editing; MI – sample collection, data collection, writing review and editing; JAD – data collection, project administration, funding acquisition, writing review and editing; AC – sample analysis, data analysis, writing review and editing; SD – study design, conceptualisation, writing review and editing; MK – sample analysis, validation, writing review and editing; JTA – sample analysis, validation, writing review and editing; EBV – sample analysis, data analysis, writing review and editing; TTT – sample analysis, data analysis, writing review and editing; JTV – sample analysis, funding acquisition, writing review and editing; AM – sample analysis, interpretation, writing review and editing; FM – sample analysis, validation, interpretation, funding acquisition, writing review and editing; PA – study design, conceptualization, interpretation, writing original draft, writing review and editing; BH – study design, conceptualization, interpretation, funding acquisition, writing review and editing; TBD – sample collection, sample analysis, writing review and editing; JCH – study design, conceptualisation, sample collection, data collection, data curation, interpretation, project administration, funding acquisition, writing original draft, writing review and editing; FL-J – study design, conceptualisation, sample collection, data collection, data analysis, project administration, writing of original draft, writing review and editing.

