## Supplemental Table 2-3 for "Breakthrough infections with the omicron and delta variants of SARS-CoV-2 result in similar re-activation of vaccine-induced immunity"

**SUPPLEMENTAL MATERIAL:**

**Supplemental Table 1:** The Excel file contains worksheets with source data for Figs 2-4 in context of vaccine status, type of infection and age. Separate sheets contain raw data for biochemical analysis and markers of inflammation and results from *in vitro* neutralization assays for SARS-CoV-2.

**Supplemental Table 2.** Clinical biochemistry at admission (T1) and 1-week follow-up (T2) in COVID-19 patients

| | | Omicron, n=52 | | Delta, n=18 | | |
|---|---|---|---|---|---|---|
| | | Median (25th,75th) | Outside ref limit | Median (25th,75th) | Outside ref limit | Ref. limits |
| Hemoglobin, g/dL | T1 | 14.5 (13.5, 15.1) | 2.0 % | 14.0 (13.1, 14.5) | 5.6 % | W: 11.7-15.3; M: 13.4-17.0 |
| | T2 | 13.5 (12.7, 14.3) | 14.6 % | 13.6 (13.3, 13.8) | 14.3 % | |
| Platelet counts, x10$^9$/L | T1 | 240 (205, 280) | 4.0 % | 234 (201, 307) | 5.6 % | 145-390 |
| | T2 | 277 (236, 323) | 5.0 % | 297 (215, 324) | 7.1 % | |
| WBC, x10$^9$/L | T1 | 5.2 (4.3, 5.9) | 2.0 % | 5.3 (4.5, 7.0) | 0.0 % | 3.5-10 |
| | T2 | 5.7 (4.9, 6.6) | 4.9 % | 6.1 (5.4, 6.7) | 0.0 % | |
| Neutrophils, x10$^9$/L | T1 | 2.6 (2.1, 3.2) | 2.0 % | 2.8 (2.3, 3.8) | 5.6 % | 1.5-7.3 |
| | T2 | 3.3 (2.6, 4.0) | 2.4 % | 3.7 (3.0, 4.2) | 0.0 % | |
| Lymphocytes, x10$^9$/L | T1 | 1.9 (1.6, 2.4) | 5.9 % | 1.9 (1.5, 2.4) | 5.6 % | 1.1-3-3 |
| | T2 | 1.8 (1.5, 2.1) | 4.9 % | 1.8 (1.5, 1.9) | 0.0 % | |
| Monocytes, x10$^9$/L | T1 | 0.4 (0.3, 0.5) | 2.0 % | 0.5 (0.4, 0.5) | 5.6 % | 0.2-0.8 |
| | T2 | 0.4 (0.4, 0.5) | 0.0 % | 0.5 (0.4, 0.6) | 0.0 % | |
| AST, U/L | T1 | 23 (20, 28) | 5.8 % | 26 (20, 29) | 5.6 % | W: 15-35; M: 15-45 |
| | T2 | 23 (19, 29) | 7.1 % | 20 (17, 25) | 0.0 % | |
| ALT, U/L | T1 | 24 (18, 38) | 0.0 % | 23 (18, 31) | 0.0 % | W: 10-45; M: 10-70 |
| | T2 | 25 (17, 38) | 4.8 % | 19 (16, 23) | 7.1 % | |
| Creatinine, µmol/L | T1 | 63 (59, 73) | 0.0 % | 66 (58, 76) | 0.0 % | W: 45-90; M: 60-105 |
| | T2 | 65 (61, 75) | 0.0 % | 69 (60, 79) | 0.0 % | |
| eGFR, mL/min/1.73m$^2$ | T1 | 113 (103, 119) | 0.0 % | 108 (100, 114) | 0.0 % | >60 |
| | T2 | 113 (102, 118) | 0.0 % | 104 (89, 111) | 0.0 % | |
| Ferritin, µg/L | T1 | 119 (66, 221) | 7.7 % | 159 (72, 201) | 5.6 % | W: 10-170; M: 30-400 |
| | T2 | 103 (65, 191) | 4.8 % | 93 (48, 235) | 21.4 % | |
| Fibrinogen, g/L | T1 | 3.3 (3.1, 4.1) | 26.8 % | 3.6 (3.2, 4.1) | 27.8 % | 1.9-4.0 |
| | T2 | 2.7 (2.4, 3.2) | 4.8 % | 2.9 (2.2, 3.5) | 0.0 % | |
| LD, U/L | T1 | 172 (160, 186) | 9.6 % | 169 (158, 178) | 0.0 % | 105-205 |
| | T2 | 172 (158, 192) | 11.9 % | 161 (146, 178) | 0.0 % | |
| ALP, U/L | T1 | 66 (51, 80) | 1.9 % | 64 (55, 79) | 5.6 % | 35-105 |
| | T2 | 60 (50, 75) | 2.4 % | 63 (49, 69) | 0.0 % | |
| CRP, mg/L | T1 | 1.8 (0.7, 4.9) | 28.8 % | 1.9 (1.0, 4.4) | 27.8 % | <4 |
| | T2 | 0.5 (0.5, 0.9) | 4.8 % | 0.6 (0.5, 1.1) | 14.3 % | |
| Creatin kinase, U/L | T1 | 76 (60, 102) | 1.9 % | 72 (51, 104) | 0.0 % | W: 35-210; |

| | | | | | | |
|---|---|---|---|---|---|---|
| | T2 | 85 (67, 111) | 2.4 % | 70 (59, 93) | 0.0 % | M: 50-400 |
| D-dimer, mg/L FEU | T1 | 0.23 (0.18, 0.32) | 7.3 % | 0.23 (0.19, 0.31) | 5.6 % | <50 yr: <0.50; >50 yr <age/100 |
| | T2 | 0.19 (0.18, 0.32) | 11.9 % | 0.22 (0.18, 0.42) | 14.3 % | |
| Troponin T, ng/L | T1 | 4 (4, 4) | 1.9 % | 4 (4, 4) | 11.1 % | <14 |
| | T2 | 4 (4, 5) | 0.0 % | 4 (4, 4) | 7.1 % | |
| NT-proBNP | T1 | 49 (49, 49) | 0.0 % | 49 (49, 63) | 0.0 % | W <50 yr: <170; >50 yr <300 |
| | T2 | 49 (49, 62) | 0.0 % | 56 (49, 80) | 0.0 % | M <50 yr: <85; >50 yr <250 |
| IgG, g/L | T1 | 11.0 (10.0, 12.3) | 3.8 % | 10.8 (9.7, 12.4) | 11.1 % | W: 6.9-15.7; M: 6.1-14-9 |
| | T2 | 10.6 (9.3, 11.5) | 0.0 % | 11.0 (9.3, 12.1) | 7.1 % | |
| IgA, g/L | T1 | 1.9 (1.65, 2.45) | 7.7 % | 2.8 (2.0, 3.1)* | 16.7 % | 0.7-3.7 |
| | T2 | 1.8 (1.5, 2.2) | 2.4 % | 2.4 (1.9, 2.9)* | 7.1 % | |
| IgM, g/L | T1 | 1.2 (0.89, 1.6) | 7.7 % | 1.35 (0.82, 1.7) | 0.0 % | 0.4-2.1 |
| | T2 | 1.1 (0.83, 1.6) | 2.4 % | 1.05 (0.76, 1.6) | 0.0 % | |

*p<0.05 vs. omicron. WBC, white blood count; AST, aspartate aminotransferase; ALT, alanine aminotransferase; eGFR, estimated glomerular filtration rate; LD, lactate dehydrogenase; ALP, alkaline phosphatase; CRP, C-reactive protein; NT-proBNP, N-terminal pro-brain natriuretic peptide; Ig, immunoglobulin.

**Supplemental Table 3:** Markers of immune activation and inflammation in COVID-19 patients and healthy controls (HC) at admission (T1) and follow-up (T2).

| | | HC, n=13 | Omicron, n=52 | Delta, n=18 | p-value |
|---|---|---|---|---|---|
| sCD14 µg/mL | T1 | 0.71 (0.44,0.82) | 1.08 (0.95,1.21)*** | 1.09 (0.95,1.32)*** | <0.001 |
| | T2 | | 0.73 (0.6,0.84) | 0.63 (0.59,0.74) | 0.12 |
| sCD163 ng/mL | T1 | 325 (301,422) | 540 (446,645)*** | 555 (489,726)*** | <0.001 |
| | T2 | | 390 (327,510)† | 340 (268,369) | 0.035 |
| MPO Ng/mL | T1 | 7.34 (5.76,11.07) | 13.3 (7.3,17.16) | 14.02 (10.51,17.91) | 0.054 |
| | T2 | | 8.27 (6,10.88) | 8.71 (5.3,14.12) | 0.93 |
| sCD25 pg/mL | T1 | 563 (459,603) | 623 (508,875) | 537 (430,708) | 0.16 |
| | T2 | | 552 (411,730) | 651 (456,705) | 0.71 |
| sTIM3 ng/mL | T1 | 5.17 (4.56,6.88) | 6.03 (5.11,7.13) | 6.12 (5.24,6.87) | 0.46 |
| | T2 | | 5.21 (4.79,6.53) | 5.57 (4.71,6.88) | 0.89 |
| LBP µg/mL | T1 | 6.48 (5.52,8.16) | 8.88 (7.36,11.23)** | 8.9 (7.15,10.59)** | 0.010 |
| | T2 | | 7.48 (5.96,8.84) | 8.73 (6.88,10.4) | 0.072 |
| TCC AU/mL | T1 | 0.87 (0.77,1.2) | 0.94 (0.72,1.17) | 0.91 (0.81,1.53) | 0.76 |
| | T2 | | 0.87 (0.67,1.09) | 0.94 (0.82,1.03) | 0.45 |
| IP10 Pg/mL | T1 | 80 (51,92) | 113 (76,281)** | 97 (71,162)* | 0.030 |
| | T2 | | 72 (45,93) | 69 (50,102) | 0.89 |
| vWF AU | T1 | 90.7 (87.9,97.2) | 72.1 (58.9,88.9)** | 79.7 (63.7,93.8)* | 0.009 |
| | T2 | | 100.7 (86.3,112.9) | 102.5 (92.2,108.8) | 0.29 |
| PTX3 Ng/mL | T1 | 1.26 (1,1.92) | 2.29 (1.63,4.3)** | 1.9 (1.52,3.51)* | 0.009 |
| | T2 | | 1.76 (1.22,2.27) | 2.39 (1.86,2.86)** | 0.016 |
| GDF15 Pg/mL | T1 | 201 (186,252) | 322 (248,442)*** | 272 (244,365)* | 0.001 |
| | T2 | | 304 (216,416)** | 300 (228,377)* | 0.010 |

| | | | | | |
|---|---|---|---|---|---|
| YKL40 Ng/mL | T1 | 31.2 (28.3,40.9) | 35.1 (28.7,40.4) | 32.4 (30.2,43.7) | 0.82 |
| | T2 | | 37.7 (31.5,47.1) | 35.6 (28.5,53.5) | 0.27 |
| MMP9 Ng/mL | T1 | 28.1 (18.4,46) | 25 (20.4,32.8) | 27.9 (20.8,43.7) | 0.83 |
| | T2 | | 27.5 (21.6,36.9) | 26.6 (17.4,41.8) | 0.97 |
| S100A12 Ng/mL | T1 | 1.94 (1.34,2.33) | 2.45 (1.69,3.85) | 2.16 (1.42,3.92) | 0.24 |
| | T2 | | 2.16 (1.84,2.93) | 1.99 (1.45,3.43) | 0.24 |

P-value in right column is from the a priori Kruskal-Wallis analysis. *p<0.05, **p<0.01, ***p<0.001 vs. HC. s, soluble; MPO, myeloperoxidase; TIM3, T cell immunoglobulin and mucin-domain containing-3; LBP, lipopolysaccharide binding protein; TCC, terminal complement complex; IP10, Interferon gamma-induced protein 10; vWF, von Willebrand factor; PTX3, Pentraxin 3; GDF15, Growth Differentiation Factor 15; YKL-40, Chitinase 3-like 1; MMP9, Matrix metallopeptidase 9; S100A12, S100 calcium-binding protein A12.
